# Efficacy of Shock Team Implementation for Cardiogenic Shock: A Systematic Review

**DOI:** 10.1101/2023.09.01.23294969

**Authors:** Mohamed Abdelnabi, Ahmed Saad Elsaeidy, Aya Moustafa Aboutaleb, Amit Johanis, Ahmed K. Ghanem, Basel Abdelazeem

## Abstract

**Introduction:** Cardiogenic shock (CS) is a critical cardiac condition characterized by low cardiac output leading to end-organ hypoperfusion and associated with high in-hospital mortality rates. It can manifest following acute myocardial infarction or acute exacerbation of chronic heart failure. Despite advancements, mortality rates remain elevated, prompting interest in multidisciplinary approaches to improve outcomes. This manuscript presents a review focused on the concept of a CS team and its potential impact on patient management and outcomes.

**Methods:** A comprehensive search was performed on March 19th, 2023, covering PubMed, Web of Science, Scopus, Embase, and Cochrane Library. We included primary studies (prospective and retrospective) only and evaluated their quality using the Newcastle-Ottawa Quality Scale. This review was registered in PROSPERO (<u>CRD42023440354</u>).

**Results:** Six relevant studies with 2066 CS patients were included, of which 1071 were managed by shock teams and 995 received standard care. Findings from the reviewed studies indicated the favorable outcomes associated with implementing CS teams. Patients managed by these teams exhibited higher 30-day and in-hospital survival rates compared to those without team intervention. The implementation of CS teams was linked to reduced in-hospital and ICU mortality rates. Additionally, shock team involvement was associated with shorter door-to-balloon times.

**Conclusion:** This review highlighted the positive influence of CS teams on patient care, enabling early detection, timely interventions, and shorter ICU stays. Despite implementation challenges, CS teams hold promise for improving management outcomes, necessitating increased attention and ongoing research in multidisciplinary strategies to advance CS care.

## 1.0 Introduction

Cardiogenic shock (CS) is a hemodynamically complex cardiac disorder associated with low cardiac output that leads to clinical manifestations and biochemical evidence of end-organ hypoperfusion. CS can present with different phenotypes, most commonly as a complication of acute myocardial infarction (AMI-CS). It can also result from different etiologies, like an acute exacerbation of chronic heart failure (non-AMI-CS). Notably, CS is associated with high in-hospital mortality ranging between 30% and 60% ^1^. AMI-CS mortality has slightly improved due to early intervention with Percutaneous Coronary Intervention (PCI) to 50.3%, as reported in the SHOCK trial ^2^.

Subsequent clinical trials have failed to show improvement in mortality with additional mechanical support for revascularization, such as Intra-aortic balloon pump (IABP) ^3–5^. However, early application of mechanical support was reported to improve mortality ^6^. Tehrani et al. reported that every 1-hour delay in MCS therapy was associated with a 9.9% increased risk of death ^7^. The improvement in outcome with the early application of MCS goes back to the pathophysiology of CS. CS progresses from a treatable hemodynamic problem to a hemo-metabolic problem that does not respond to the MCS. Knowing that the concept of “door to support” time started to gain more attention in treating CS ^8^. Early identification of CS can be challenging because patients can present to the ER or the health care facility in different stages and phenotypes. In other critical health conditions like pulmonary embolism and stroke, early identification and intervention by a dedicated rapid response team were associated with a marked decrease in mortality and morbidity ^9,10^. From here, the idea of creating a multidisciplinary team for the early identification and management of CS started to be implemented. Different centers applied this concept and found that managing CS with a rapid-response multidisciplinary team was associated with decreased mortality ^7,11–16^. In this review, we will discuss the concept of a CS shock team and the outcomes of its implementation.

## 2.0 Materials and Methods

### 2.1. Search strategy

We searched PubMed, Web of Science, Scopus, Embase, and Cochrane Center till March 19^th^, 2023. We used the following search strategy to find all the studies discussing the management of cardiogenic shock with versus without shock teams (“ECMO team” OR “Multidisciplinary Care Team” OR “Interdisciplinary Health Team” OR “Shock team” OR “Rapid Response Team”) AND (“Cardiac shock” OR “Cardiogenic Shock”) as shown in the appendix. We gathered the search terms from the MeSH database and the literature and then built the strategy as described in the Cochrane Handbook for Systematic Reviews of Interventions (Chapter 4.4.4) ^17^.

Our review was performed in accordance with the Preferred Reporting Items for Systematic Reviews and Meta-Analyses (PRISMA) statement ^18^. This review was registered in PROSPERO (<u>CRD42023440354</u>).

### 2.2. Selection Criteria and Screening

We included primary studies (prospective and retrospective) published in peer-reviewed journals comparing critical outcomes of cardiogenic shock in adult patients who were managed with shock teams versus those without shock teams. We excluded animal studies, case reports, letters to the editor, conference abstracts, and secondary studies. Three authors (A.S.E, A.M.A, M.A) screened the articles by title and abstract, then by reading the full texts using Covidence systematic review software (Available at www.covidence.org).

### 2.3. Data Extraction

We extracted data summarizing the included studies’ criteria, demographics, and baseline characteristics of their patients, including CS etiology, cardiac arrest, baseline lab values, and cardiovascular risks.

Additionally, we extracted the studies’ outcome data, including the rates of 30-day survival, in-hospital survival, in-hospital mortality, ICU mortality, time to treatment (door-to-balloon time), rate of MCS utilization, and the use of mechanical ventilation and renal replacement therapy. Two authors (A.S.E, A.M.A) extracted the data and then all the extracted data were revised by a third author (M.A).

### 2.4. Quality Assessment

Two authors (A.S.E, A.M.A) evaluated the quality of the included studies using the Newcastle-Ottawa Quality Scale (NOS) ^19^. The NOS evaluates the studies’ quality according to three domains: selection of study population; comparability between study cases and controls; and exposure determination. The results from the NOS were converted into the Agency for Healthcare Research and Quality (AHRQ) standards as the following: A) Good quality: 3 or 4 stars in selection domain, 1 or 2 stars in comparability domain, and 2 or 3 stars in outcome/exposure domain; B) Fair quality: 2 stars in selection domain, 1 or 2 stars in comparability domain, and 2 or 3 stars in outcome/exposure domain; C) Poor quality: 0 or 1 star in selection domain, 0 stars in comparability domain, or 0 or 1 star in outcome/exposure domain ^20^. According to AHRQ standards, two of the included studies in our review have good quality ^11,13^, and four studies have poor quality ^7,12,14,15^, as shown in Table 3.

## 3.0 Results

### 3.1. Search results

The search strategy over the different medical databases yielded 1683 after removing duplicates. Screening the title and the abstract yielded 23 studies after excluding 1660 papers because they were animal studies, out of the study criteria, and/or not primary studies. Twenty-three articles were screened for their full-text testing for eligibility. Six papers met the study criteria ^7,11–15^ and were further included in the quality assessment, as shown in Fig.1.

**Figure 1:**
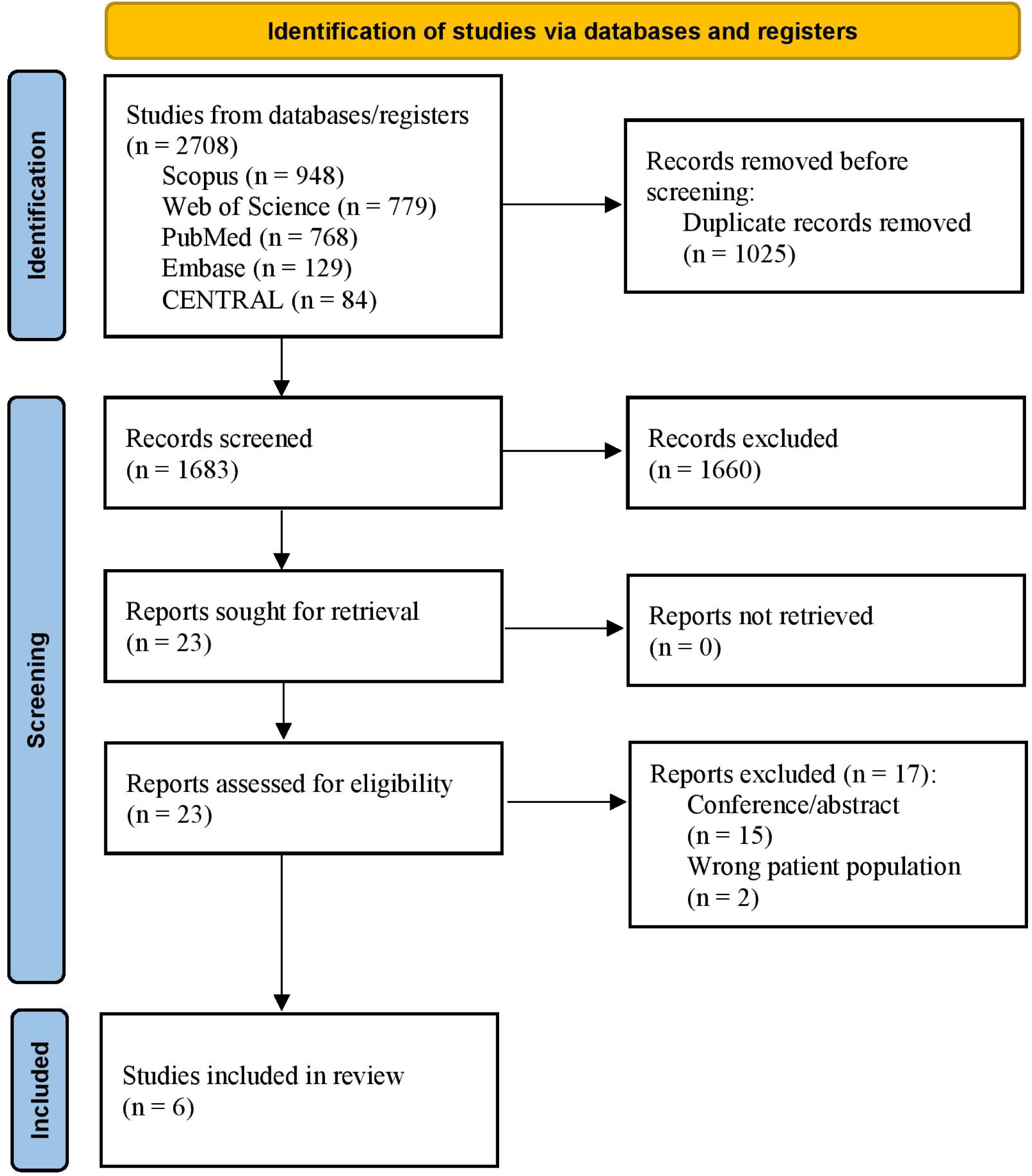
PRISMA flowchart of the database search and searching process.

### 3.2. Baseline Characteristics and Outcomes

All the included studies are peer-reviewed cohort studies. Three of the studies are retrospective cohort studies ^13–15^, while the other three are retrospective-prospective cohorts. Five studies are from the USA ^7,11,12,14,15^ or Canada, and one study is from Korea ^13^. The included studies encompass data from 2066 cardiogenic shock patients, with 1071 patients being treated by the shock team and 995 without the shock team’s intervention. The mean age of the participants is greater than 50 years old. These studies consist of 1433 male patients, 850 patients with acute myocardial infarction-related cardiogenic shock, 1195 patients with non-AMI CS, 541 patients who experienced cardiac arrest, and 293 patients with diabetes mellitus. Further details regarding baseline characteristics and baseline laboratory work, as well as cardiovascular risks, are summarized in Table 1.

**Table 1.** Baseline characteristics.

| Table 1. Baseline characteristics. |  |  |  |  |  |  |  |  |  |  |  |  |  |
| --- | --- | --- | --- | --- | --- | --- | --- | --- | --- | --- | --- | --- | --- |
| study ID | Group | Demographics |  |  | Cardiogenic shock etiology |  | Cardiac Arrest |  | Baseline lab. |  | Cardiovascular risks |  |  |
|  |  | Sample size | Age, y, M(SD) | Male, n (%) | AMI-CS, n (%) | Non-AMI-CS, n (%) | Out-of-hospital, n (%) | In-hospital, n (%) | Baseline Creatinine, mg/dL, M(SD) | Baseline Lactic acid, mmol/L, M(SD) | Diabetes mellitus, n (%) | Baseline cardiac index, L/min/m2, M(SD) | Left ventricular ejection fraction, M(SD) |
| Hong et al. 2020 | CS team | 124 | 64.7 (11.5) | 95 (76.6) | 124 (100%) | 0 | 22 (17.7) | 81 (65.3) | 1.3 (0.5) | 5.03 (4.05) | 61 (49.2%) | NA | 31.5 (11.63) |
|  | Control | 131 | 63.4 (12) | 105 (80.2) | 131 (100%) | 0 | 17 (13) | 82 (62.6) | 1.3 (0.6) | 6.5 (5.7) | 67 (51.1%) |  | 34 (14.99) |
| Lee et al. 2020 | CS team | 64 | 53.9 (15.8) | 50 (78) | 7 (11) | 57 (89) | 13 (20) |  | 1.5 (0.7) | 3.6 (2.6) | NA | 1.65 (0.6) | 18.3 (9.9) |
|  | Control | 36 | 60.1 (16.1) | 24 (67) | 6 (17) | 30 (83) | 8 (22) |  | 2 (1.5) | 3.1 (2.2) |  | 2.03 (1) | 21.7 (11.6) |
| Papolos et al 2021 | CS team | 546 | 64.7 (15.6) | 351 (64.3) | 147 (26.9) | 399 (73.1) | 61 (12) | 67 (13.1) | 1.6 (0.9) | 3.77 (2.38) | NA | 1.9 (0.54) | **CS / control**<br>≥50: 87 (17.09) / 88 (13.79)<br>20-50: 267 (52.46) / 255 (39.97)<br><20: 55 (30.45%) / 295 (46.24) |
|  | Control | 696 | 63 (13.4) | 477 (68.5) | 193 (27.7) | 503 (72.3) | 77 (11.8) | 83 (12.7) | 1.7 (0.9) | 2.7 (2.23) |  | 2.04 (0.67) |  |
| Taleb et al. 2019 | CS team | 123 | 57 (1) | 96 (78.04) | 75 (60.97) | 48 (39.02) | NA | NA | NA | 5.4 (0.5) | 37 (30.08) | 2.2 (0.2) | 24 (2) |
|  | Control | 121 | 59 (1) | 93 (76.85) | 85 (70.25) | 36 (29.75) |  |  |  | 5.9 (0.5) | 35 (28.92) | 2.2 (0.2) | 28 (2) |
| Tehrani et al. 2019 | AMI | 82 | 64 (11) | 58 (70.73) | 82 (100) | 0 (0) | 9 (10.98) | 10 (12.2) | NA | 4.9 (4.3) | 46 (56.09) | <1.8 / <2.2 l/min/m2 without / with inotropes/vasopressors | NA |
|  | Decompensated HF | 122 | 58.4 (14) | 84 (68.85) | 0 (0) | 122 (100) | 6 (4.92) | 5 (4.1) |  | 4.3 (3.8) | 47 (38.52) |  |  |
|  | Control | NA |  |  |  |  |  |  |  | NA |  |  |  |
| Sebat et al. 2005 | CS team | 11 | NA | NA | NA | NA | NA | NA | NA | NA | NA | ≥ 2.5 L/min/m2 | NA |
|  | Control | 10 |  |  |  |  |  |  |  |  |  |  |  |

The data are reported as n (%) or mean ± standard deviation.

Sebat et al. excluded all patients who were not eligible for aggressive treatment or were suffering from non-survivable conditions that preceded the shock state, such as untreatable metastatic carcinoma, brain death, or ruptured thoracic aneurysm ^12^. Similarly, Lee et al. excluded terminally-ill patients with a life expectancy of less than six months, as well as those who experienced cardiac arrest for more than 30 minutes ^14^. While Hong et al. included patients with AMI-CS undergoing veno-arterial extracorporeal membrane oxygenation (VA-ECMO), they excluded stable patients who received prophylactic VA-ECMO before revascularization ^13^. Taleb et al. also excluded postcardiotomy patients and those who required ECMO ^11^.

Three studies reported the 30-day survival rate, two of them (Hong et al. & Taleb et al.) found significantly higher survival rates among CS patients treated with the shock team ^11,13,14^. Two studies investigated the in-hospital survival rate, one of them (Taleb et al.) reported a higher survival rate in the CS team group ^11^, while the other (Lee et al.) reported no significant difference between both groups ^14^. Hong et al. and Sebat et al. investigated the in-hospital mortality rate and reported lower rates with the implementation of the shock team ^13,12^. Hong et al. and Papolos et al. reported the ICU mortality rate and found a lower mortality rate for the patients treated with shock team ^13,15^. Also, Hong et al. and Taleb et al. reported door-to-balloon time and found a shorter time to start treating patients with shock team ^11,13^. However, Hong et al. reported that NSTEMI Shock patients treated with a shock team spent more time to start the treatment ^13^. Moreover, the utilization rate of mechanical circulatory support (MCS) was documented in studies by Lee et al. and Papolos et al. The former demonstrated a higher rate of MCS utilization in the CS team group, whereas the latter presented contrasting results ^14,15^.

Additionally, we summarized more details about the reported measures for CS patients’ management, such as mechanical ventilation, and renal replacement therapy in Table 2.

**Table 2.** Studies summary, Outcomes, and treatment.

The data are reported as n (%) or mean ± standard deviation.
| Table 2. Studies summary, Outcomes, and treatment. |  |  |  |  |  |  |  |  |  |  |  |  |
| --- | --- | --- | --- | --- | --- | --- | --- | --- | --- | --- | --- | --- |
| Study ID | Country | Study design | Group | no. of patients | 30-day survival, n(%) | In hospital survival, n(%) | In hospital mortality, n(%) | ICU mortality, n(%) | time to treatment(door-to-balloon time), min, M(SD) | rate of MCS utilization, n(%) | Mechanical ventilation n(%) | Renal replacement therapy n(%) |
| Hong et al. 2020 | Republic of Korea | Retrospective cohort study | CS team | 124 | 83 (66.9) | NA | 42 (33.9) | 38 (30.6) | -STEMI: 90.5 (34.13)<br>-NSTEMI: 918 (1530.16) | NA | 102 (82.3) | Continuous RRT 50 (40.3) |
|  |  |  | Control | 131 | 58 (44.3) |  | 71 (54.2) | 68 (51.9) | - STEMI: 114.33(62.22)<br>- NSTEMI: 389.33(478.24) |  | 124 (94.7) | 49 (37.4) |
| Lee et al. 2020 | Canada | Retrospective cohort study | CS team | 64 | 46 (72) | 44 (69) | NA | NA | NA | 29 (45) | Invasive ventilation: 41(64) | Dialysis:16 (25) |
|  |  |  | Control | 36 | 25 (69) | 22 (61) |  |  |  | 10 (28) | Invasive ventilation: 25(69) | Dialysis: 17 (47) |
| Papolos et al. 2021 | United States and Canada | Retrospective cohort study | CS team | 546 | NA | NA | NA | 126 (23.1) | NA | 192 (35.2) | 223 (40.8) | New RRT: 58(10.6) |
|  |  |  | Control | 696 |  |  |  | 200 (28.7) |  | 299 (43.0) | 363 (52.2) | New RRT: 131(18.8) |
| Taleb et al. 2019 | United States | Retrospective and prospective cohort study | CS team | 123 | 88 (71.85) | 75 (61.0) | NA | NA | 19±5 h | NA | 81 (65.9) | NA |
|  |  | (control cohort) | Control | 121 | 67 (55.56) | 58 (47.9) |  |  | 25±8 h |  | 81 (66.9) |  |
| Tehrani et al. 2019 | United States | Retrospective and prospective cohort study (control cohort) | AMI | 82 | 52 (63.4) | NA | NA | NA | NA | NA | NA | Dialysis:27(32.9)<br>Transplant: 0(0.0) |
|  |  |  | Acute Decompensated HF (ADHF) | 122 | 78(63.9) |  |  |  |  |  |  | Dialysis: 34(27.9)<br>Transplant: 5(4.1) |
|  |  |  | Control | NA | NA(47) |  |  |  |  |  |  | NA |
| Sebat et al. 2005 | United States | Retrospective and prospective cohort study (control cohort) | CS team | 10 | NA | NA | 2 (0.2) | NA | NA | NA | NA | NA |
|  |  |  | Control | 11 |  |  | 4 (36.36) |  |  |  |  |  |

## 4.0 Discussion

### 4.1. Definition of CS

Early identification and diagnosis of cardiogenic shock are critical to improving its poor outcomes. CS is pragmatically referred to as the state of impaired cardiac output and decreasing tissue perfusion. The first definition of CS was mentioned in the SHOCK trial ^2^ based on clinical and hemodynamic indices. Clinical indices include systolic blood pressure (SBP) <90 mmHg for ≥30 min OR SBP ≥90 mmHg with support AND end-organ hypoperfusion (i.e. urine output (UOP) <30 mL/h or cool extremities). Hemodynamic indices include a Cardiac Index (CI) of ≤2.2 L/min/m2 AND pulmonary capillary wedge pressure (PCWP) ≥15 mmHg. The IABP-SHOCK II trial ^3^ defined CS as the following: SBP <90 mmHg for ≥30 min OR catecholamines to maintain SBP >90 mmHg AND clinical pulmonary congestion AND impaired end-organ perfusion including altered mental status, cold/clammy skin, and extremities, UOP <30 mL/h, or lactate >2.0 mmol/L. A recent definition of CS was mentioned by the European Society of Cardiology (ESC) ^21^ as SBP <90 mmHg with adequate volume AND clinical signs of hypoperfusion, including cold extremities, oliguria, mental confusion, dizziness, narrow pulse pressure, OR laboratory signs of hypoperfusion including metabolic acidosis, elevated serum lactate, and elevated serum creatinine. The Society for Cardiovascular Angiography and Intervention (SCAI) also proposed and validated a recent classification of CS into five stages ranging from ‘at risk’ to ‘extremis’ and defined CS according to the stage. For example, classic CS was described as a manifestation of hypoperfusion requiring medical or MCS intervention to restore perfusion AND relative hypotension ^43,44^.

In our review, Hong et al. used the ECS definition of CS to identify their inclusion criteria ^13^, Lee et al. and Tehrani et al. used the SHOCK trial definition ^14,16^, Lee et al. and Papolos et al. reported that most of their patient population fell into categories C “classic CS” and D “deteriorating” respectively according to the SCAI classification system ^14,15^, while Taleb et al. and Sebat et al. didn’t report the criteria based on which CS was confirmed ^11,12^.

### 4.2. Phenotypes of CS

Earlier, we mentioned that CS presents at different stages and phenotypes. The most common phenotype presents with low CI, elevated systemic vascular resistance (SVR), and high PCWP and is referred to as “cold and wet” CS ^22^. Cold and wet CS is mainly AMI-CS. Another less common phenotype is called “cold and dry”. It presents with low CI, lower PCWP, and euvolemia. Cold and dry CS is mainly non-AMI CS. A third AMI-CS phenotype is called “wet and warm”. This wet and warm CS presents with low CI, systemic inflammation, increased PCWP, and low SVR; it also has a higher risk of mortality ^23^. A fourth AMI-CS phenotype was reported in the SHOCK trial with SBP >90 mmHg without vasopressors ^24^. The last AMI-CS phenotype reported in the SHOCK trial is right ventricular (RV) CS ^25^. Heart Failure (HF)-CS is another phenotype which relates to CS that develops as a complication of acute decompensated HF (ADHF), and is also mostly described as “wet and warm” ^45^. Overall, the common characteristic between all phenotypes is low CI.

### 4.3. Multidisciplinary CS team

Given the different phenotypes, complexity, and high mortality of CS, new approaches to improving its outcomes started to be embraced. A cardiogenic shock team, or shock team, is one of the newly recognized approaches for managing CS and has shown favorable outcomes. The idea of creating a shock team was inspired by other teams developed to manage acute complex health problems, such as stroke teams and ST-elevation myocardial infarction (STEMI) teams. The shock team is a multidisciplinary team that gets activated to identify and manage patients who meet predefined criteria for CS. To date, there is no unified structure for the shock team. However, the most frequently reported members of the team in the studies included in our review are a cardiothoracic surgeon, an interventional cardiologist, a heart failure specialist, a critical care cardiologist, and an emergency physician. These members cover four main areas, including the intensive care unit (ICU), cardiac catheterization lab, cardiac surgery, and advanced heart failure ^26–28^. Support from trained nurses, perfusionists, and respiratory therapists is also required. The team has a coordinating physician or a team leader who gets notified first once a suspected CS patient presents. The team leader, in turn, notifies the rest of the team members and activates the team as well as the supporting staff. Most of the time, the team leader is the on-call critical care cardiologist. To make the team more efficient, different departments should be aware of the shock team and the adopted local guidelines for CS identification and thus notify the team leader whenever a case of CS is suspected. The team leader responds to these notifications, assesses the case, and then activates the team ^28^. If the shock is AMI-CS, the interventional cardiologist and the cardiothoracic surgeon start the revascularization procedures and provide large-bore vascular access for MCS and emergency cardiac surgeries as needed. The heart failure specialist and critical care cardiologists usually continue to care for the patients after the interventional procedures. For patients presenting with non-AMI CS, the critical care cardiologist, and heart failure specialist initiate a complete assessment with the interventional team members on standby if needed.

Although shock team members may have overlapping medical skills, every member has a unique skill that adds to the team’s ability to maximize the patient’s outcome. Therefore, all team members should be actively involved in decision-making and considerate of the long-term plan for patient management.

### 4.4. Multidisciplinary CS-team and clinical outcomes

CS patients treated with CS team showed an overall higher 30-day survival rate, reduced in-hospital and ICU mortality, shorter door-to-balloon time, faster initiation of therapies, including MCS and less need for mechanical ventilation and renal replacement therapy.

#### 4.4.1. 30-day Survival

Tehrani et al. looked at the outcomes of implementing a CS “shock team” for treating patients with CS. Their study consisted of 204 patients with CS who were monitored for 18 months. Notably, the 30-day survival of CS patients post-discharge increased from a baseline survival of 47% to 58% in the first year and 77% in the second year after the implementation of a shock team, and to 52% and 78% in AMI and ADHF patients respectively (Table 2) ^16^.

Similarly, Taleb et al. also compared the outcomes of treating CS with and without designated CS teams. They found a decrease in 30-day all-cause mortality for CS patients treated with the shock team (Table 2).

#### 4.4.2. In-hospital Survival

Taleb et al. found an increase in in-hospital survival (61% vs. 47%) for patients treated by a shock team when compared with non-shock team (Table 2). Contrarily, Lee et al. reported no statistically significant difference in the short-term survival rates between the CS patients treated with shock team vs. control team. However, the overall survival showed an improvement of 67% over a follow-up duration of 240 days. These findings support the efficacy of implementing CS teams, and especially noteworthy are the profound improvements in survival ^11^.

#### 4.4.3. In-hospital Mortality

Hong et al. reported significantly reduced in-hospital mortality (71% vs. 42%) among CS patients in the Shock team group vs. control group ^13^. Sebat et al. also reported an improvement of in-hospital mortality with the shock team implementation despite the small population of CS patients included in their study (Table 2) ^12^.

#### 4.4.4. ICU Mortality

Papolos et al. reported that the duration of stay in a cardiac intensive care unit (CICU) and CS outcomes showed considerable improvements. Patients under the care of a CS team were in the CICU for an average of 4 days, compared to 5.1 days for patients treated without a designated CS team. Furthermore, there was a reduction in mortality for patients in CICUs with a CS team compared to those in CICUs without a CS team (23% vs 29% respectively) (Table 2) ^15^.

Similarly, another study conducted by Hong et al. compared mortality rates for 255 AMI-CS patients before and after the development of a designated multidisciplinary team. They found a significant decrease in in-hospital mortality (54% vs. 33%) and CICU mortality (45% vs. 25%) after implementation and treatment by a multidisciplinary ECMO team (Table 2). Furthermore, the study also found a decrease in both all-cause mortality (58% vs. 35%) and readmission rates for heart failure (28% vs. 6%) under an ECMO team at a 6-month follow-up ^13^.

Likewise, a study by Lee et al. found improved outcomes after the implementation of a multidisciplinary ECMO team for patients who underwent in-hospital cardiac arrest ^14^. Specifically, their study showed decreases in both in-hospital mortality (75% vs. 40%) and negative neurological outcomes (78% vs. 48%) for these patients after the implementation of the ECMO team. Interestingly, there was no significant difference in either in-hospital mortality or neurological outcomes for patients who experienced cardiac arrest outside of the hospital, regardless of whether the ECMO team was utilized or not. Lee et al. reason that during a cardiac arrest, it is often difficult to determine appropriate interventions, especially due to a lack of information in a real-time situation; hence, a multidisciplinary team of experts can help determine optimal strategies and timing for advanced support, such as ECMO to help improve outcomes in critically acute situations ^14^.

Lastly, no differences were found in ICU stay or rates of major complications such as bleeding, cerebrovascular events, hemolysis, or other vascular complications between the experimental or control groups ^11^. This could be an area for further research, specifically targeting the management of complications related to CS.

#### 4.4.5. Time to treatment (Door-to-balloon time)

Along with the deleterious effects that can be attributed to delays in identification and intervention for CS, another important factor to consider is the time required for stabilizing a very sick patient with CS ^26^. In the case of AMI-CS, less than 40% of these patients are treated within the recommended contact-to-device time (90 minutes) ^29^. Furthermore, findings by Scholz et al. have demonstrated that a 10-minute delay in primary PCI for a patient presenting with CS led to over three additional deaths out of every 100 patients treated with PCI ^30^. Moreover, the time between the initial presentation of a CS patient and PCI intervention has a strong association with a negative outcome. As a result, door-to-support time became an emergent concept for managing AMI-CS. This is important to consider as studies have demonstrated that the timely deployment of MCS devices have led to better clinical outcomes ^31,32^. Providing early and effective MCS, especially in AMI-CS, helps unload the left ventricle, prevents or can even reverse end-organ damage, decreases myocardial wall tension, and, in turn, improves outcomes ^8,33^. Basir et al. reported that early implementation of MCS in patients with cardiogenic shock independently improved survival^6^. When MCS was initiated less than 1.25 hours from shock onset, the survival of patients with CS was 66%, when initiated within 1.25 to 4.25 hours, survival was 37%, and 26% when initiated after 4.25 hours. Papolos et al. compared the outcomes of managing CS patients with shock teams versus without and found that facilities with a shock team had increased pulmonary arterial catheter use (60% vs. 49%) that were initiated in nearly half the time (0.3 days vs. 0.66 days) ^15^. Furthermore, both Hong et al. and Sebat et al. reported a shorter door-to-balloon (door-to-support) time after implementing a multidisciplinary team approach for treating patients with STEMI ^12,13^. Sebat et al. also reported a significant reduction in the median time for interventions in patients with CS after adopting a multidisciplinary team approach ^12^. The reported times in the CS team group were as follows: intensivist arrival, 2:00 h to 50 min (p < 0.002); ICU/operating room admission, 2 h 47 min to 1 h 30 min (p < 0.002); 2 L fluid infused, 3 h 52 min to 1 h 45 min (p < 0.0001); and pulmonary artery catheter placement, 3 h 50 min to 2 h 10 min (p 0.02). These shorter times for intervention were also associated with improved clinical outcomes, including mortality (28.2% in the CS team vs. 40.7% in the control group). Tehrani et al. reported that the decrease of 5 and 10 hours in the time to implement MCS was associated with increased survival of 53.6% and 135.8%, respectively ^16^. On the contrary, only Taleb et al. reported similar time-to-support for the CS team vs. the non-CS team ^11^. This is still important to consider as it demonstrates there are no delays in the delivery of care when utilizing multiple experts within a shock team. Hence, the majority of studies we reviewed have reported a shorter time to support when applying a multidisciplinary team approach for treating CS.

#### 4.4.6. Management of CS (medical & interventional)

Not only is the time-to-intervention by the shock team shorter, but the utilization of resources is also more efficient. Higher doses of vasopressors and delayed escalation of patients from medical treatment to more invasive interventions are associated with unfavorable outcomes ^34,35^. The short-term stabilizing effects of inotropes and vasopressors are countered by their adverse effects on end-organ hypoperfusion ^36^. The CS team optimizes the use of these pharmacological agents with consideration for the use of MCS. Papolos et al. reported that the CS team used fewer inotropic agents and MCS (35% vs. 43%) yet utilized more advanced MCS (i.e., Impella, ECMO, ventricular assist devices) as their first line when MCS was needed ^15^. Similarly, Tehrani et al. outlined a goal in their protocol adopted by the CS team to lower the use of vasopressors and inotropes, but increase the early use of MCS of the left ventricle and/or right ventricle as appropriate ^7^. Lee et al. also reported similar usage of inotropes before and after the implementation of the CS team. However, the use of MCS was higher after the implementation of the team ^14^. Hong et al. reported that the use of inotropes and vasopressors was lessened after the CS team’s implementation ^13^.

The decision to start invasive interventions is taken in a timely manner given the team’s structure and resources, which usually reflect positively on the outcomes. Tehrani et al. reported that every 1-hour delay in therapy escalation in patients requiring MCS was associated with a 9.9% increased risk of death ^7^. Starting pulmonary artery catheterization (PAC), also known as right heart catheterization (RHC), for hemodynamic monitoring can be beneficial for diagnosing and determining the severity and phenotype of the CS; this can be especially helpful in patients who are not responding to the initial therapy ^23^. This also facilitates individualized, invasive therapy tailored to the patient’s unique presentation. Despite the lack of evidence supporting the use of PAC monitoring for all patients, many studies have shown a decrease in mortality with the use of PAC monitoring in CS patients ^37,38^. Tehrani et al. showed that using RHC was associated with a decrease in 30-day mortality with an odds ratio of 0.19 (0.09–0.40) and P<0.01 ^7^. The use of RHC was also reported more frequently by the CS team in the study reported by Lee et al. ^14^. Despite the reported increase in the usage of PAC by Papolos et al., the overall usage of MCS was lower after the implementation of the CS team ^15^. Additionally, the formation of CS teams appears to influence MCS decisions. For example, the usage of Impella increased after the implementation of CS Teams ^11,14^. Also, the use of IABP was noticed to be less after the implementation of CS Teams ^11,13,15^. The type of MCS used by different teams might differ, but the rapid use and escalation of the MCS are noticed in all CS teams. MCS may also need to be initiated outside the tertiary CS centers for critical patients in a timely manner, which supports the idea of a “Hub and Spoke” model.

Furthermore, Papolos et al. reported only 41% of CS patients were treated with mechanical ventilation, and 11% required additional renal replacement therapy, compared to 52% and 19%, respectively, for patients without a CS team ^15^. Hong et al. also reported less patients in the shock team group who needed respiratory support through mechanical ventilation than without the shock team (Table 2). This might be as a result of the improved oxygenation and end-organ perfusion status of CS patients as a result of the time-critical shock team management strategies, which also involve better utilization of the resources.

### 4.5. Hub and spoke

One approach that can be implemented is the “Hub and Spoke” model to improve the response to CS cases. Specifically, it is important to address the poor mortality rates for CS patients that is often worsened due to a lack of resources. Moreover, delays in diagnosis and mobilization of resources impede prompt management and intervention ^26^. This is especially true in rural medical centers that may lack PCI and MCS capabilities, making the already underwhelming outcomes even worse ^39,40^. Hence, a “Hub” level I center that is comprised of a cardiogenic shock team with full capabilities of managing CS in a timely and efficient manner will lead to earlier access to PCI, coronary artery bypass graft (CABG), and MCS, along with expertise in the management of hemodynamics ^39^. Moreover, studies have shown that high-volume specialized and larger academic centers have better outcomes than lower-volume, smaller centers ^40^. This supports the implementation of specialized centers (hubs) and cardiogenic shock teams that are experienced in performing CS interventions and managing critically ill patients. Furthermore, this team-based model will promote open communication amongst providers, which is beneficial for appropriate patient selection and for escalating or de-escalating care for patients ^28^.

Although none of the studies included in our review discussed a potential “hub and spoke” model to follow, possibly due to the novelty of the concept, as well as the challenge of providing specifics of the model, which can vary widely based on the healthcare system and the region’s resources, SCAI have proposed a model based on the cardiogenic shock stage in their classification system. This model advises medical centers to identify as hubs or spokes according to their resources. Hubs would allow accepting CS patients from spokes at stage D “deteriorating” before further worsening into the most complex stage E “extremis” ^44^. Further research/clinical studies is warranted to propose more models, and validate the current ones, as well as discussing their applicability and potential challenges.

### 4.6. Challenges facing multidisciplinary team application

According to the Critical Care Cardiology Trials Network, 14 out of 24 centers in North America do not have shock teams ^15^. Major hurdles for incorporating a multidisciplinary team include a lack of resources that can range from staffing, education, and equipment. From the papers we have discussed, most of the cardiogenic shock teams include cross-collaboration among physicians from different specialties and subspecialties. This can be challenging as it requires a tertiary center that is well-funded and staffed, along with regular training to ensure the team is competent in dealing with diverse cases of cardiogenic shock ^26^. By that same note, education to identify and manage CS is important for both physicians and non-physician personnel, yet it can also pose a challenge. In fact, a key takeaway from Sebat et al. were the improvements in shock outcomes after implementing education for non-physician personnel, which led to earlier interventions, prevention of multi-organ damage, and improved outcomes^12^.

Furthermore, it is important to consider geographic differences in CS team implementation. Loccoh et al. studied the differences between rural and urban settings when addressing acute cardiac conditions (i.e., AMI and Heart Failure), and found that patients in rural areas underwent fewer medical procedures and had higher mortality rates (30-day and 90-day) compared to patients in urban settings ^41^. We believe this highlights the additional hurdles that underserved and rural communities will face; namely, disparities in available specialists, services offered, modernized equipment, and under-resourced facilities will all pose a challenge in implementing a CS team ^41,42^.

Additionally, Moghaddam et al. raised an important concern regarding expediency. In situations such as CS, where timing is imperative while making clinical decisions and interventions, any delays such as that relating to the involvement of multiple specialists can become detrimental if not organized and coordinated ^26^. Considering these multiple factors, we believe that the expansion of CS teams remains a challenge, especially for healthcare centers in under-resourced areas.

### 4.7. Strengths

Our systematic review discusses the most up-to-date evidence regarding the definitions and phenotypes of CS, the efficacy of the shock team implementation particularly on CS outcomes, and the potential challenges facing the shock team application.

### 4.8. Limitations

Despite the promising results, our study has several limitations. Firstly, most of the evidence provided by the included studies in our systematic review is retrospective/observational, which increases the risk of bias. Secondly, the quality of evidence according to the Newcastle-Ottawa scale for cohort studies was highlighted as “poor quality” in four of the six studies included in our review (Table 3). Thirdly, the included studies exhibited variations in the reported clinical outcomes. Additionally, some of the clinical outcome data/numbers were not reported by the studies’ investigators. This has limited our ability to conduct a meta-analysis of the available evidence due to its insufficiency. Finally, the population size reported by some of the studies in our review is small, which impacts the representativeness and generalizability of our findings.

**Table 3.** Quality assessment using the Newcastle-Ottawa scale for cohort studies.

| Study ID | Year | Selection |  |  |  | Comparability |  | Exposure |  |  | AHRQ standards |
| --- | --- | --- | --- | --- | --- | --- | --- | --- | --- | --- | --- |
|  |  | Representativeness of the exposed cohort | Selection of the non-exposed cohort | Ascertainment of exposure | Demonstration that outcome of interest was not present at the start of the study | Study controls for age | Study controls for any additional factor | Assessment of outcome | Was follow-up long enough for outcomes to occur | Adequacy of follow up of cohorts |  |
| Papolos et al | 2021 | ★ | ★ | ★ | ★ | - | - | ★ | ★ | ★ | Poor quality |
| Hong et al | 2020 | ★ | ★ | ★ | ★ | ★ | - | ★ | ★ | ★ | Good quality |
| Lee et al | 2020 | ★ | ★ | ★ | ★ | - | - | ★ | ★ | ★ | Poor quality |
| Taleb et al | 2019 | ★ | ★ | ★ | ★ | ★ | - | ★ | ★ | ★ | Good quality |
| Tehrani et al | 2019 | ★ | ★ | ★ | ★ | - | - | ★ | ★ | ★ | Poor quality |
| Sebat et al | 2005 | ★ | ★ | ★ | ★ | ★ | - | ★ | - | - | Poor quality |

### 4.9. Implications for Future Research

Our review offers valuable insights for clinicians regarding the multifaceted roles, benefits, and challenges associated with CS teams in enhancing patient outcomes. Nevertheless, there are several key implications for future research that can significantly enhance the quality of evidence. Firstly, it is advisable to conduct a greater number of prospective cohort studies and randomized controlled trials (RCTs), involving larger participant populations. These endeavors will yield more compelling evidence of the team’s effectiveness by minimizing potential confounding factors and selection biases.

Secondly, delving into the extended impact of CS teams’ interventions on the long-term outcomes of CS patients will be invaluable. This exploration can provide a comprehensive understanding of the team’s influence on factors such as patient survival, quality of life, and readmission rates.

Thirdly, it is pertinent to assess the cost-effectiveness of establishing and maintaining CS teams, as these insights can be pivotal in guiding strategic decisions within hospital management.

Lastly, it is worth noting that the current body of evidence concerning the impact of CS teams on outcomes specifically related to Acute Decompensated Heart Failure with Cardiogenic Shock (ADHF-CS) patients is notably limited. Recent research has highlighted a rising prevalence of ADHF-CS when compared to cases of AMI-CS ^46^. Consequently, further research in this realm is imperative to enhance our understanding of the distinct implications and benefits of CS teams for ADHF-CS patients.

## 5.0 Conclusion

For several years, mortality rates for CS have remained unacceptably high and have largely plateaued despite advancements in cardiac care. Aside from early PCI interventions, there have not been significant therapies to improve survival rates. However, a number of studies in our review have demonstrated significant reductions in both in-hospital and all-cause mortality after the addition of cardiogenic shock teams. Moreover, it is becoming evident that multidisciplinary teams are indispensable for improving clinical outcomes in cardiogenic shock cases. These studies showed additional benefits, including earlier diagnosis of CS, rapid initiation of appropriate therapies, including MCS when required, reduced ICU stays, and overall expertise in treating a broad spectrum of CS patients. That being said, there are many challenges involved with organizing a well-trained CS team to optimize clinical outcomes, and we understand that this will be an even greater challenge for underserved and rural facilities to implement. Additionally, there is a lack of research on the efficacy of a multidisciplinary approach for managing CS complications, and we find this to be a valuable area for further research. Overall, we are hopeful that emerging evidence demonstrating the benefits of CS teams will help shift momentum toward CS team implementation and improve care for CS patients.

## DECLARATION

### Ethics approval and consent to participate

Not applicable.

### Consent for publication

Not applicable.

### Availability of data and materials

Data will be provided upon request from Ahmed Saad Elsaeidy.

## Funding

This research received no specific grant from funding agencies in the public, commercial, or not-for-profit sectors.

### Authors’ contributions

**M.A, B.A, A.S.E, and A.M.A:** Conceptualization, Methodology. **A.S.E, A.MA, A.J, and A.G:** Data curation, Writing-Original draft preparation. **A.S.E and A.M.A:** Visualization, Investigation. **M.A and B.A:** Supervision. **M.A and B.A:** Software, Validation. **A.S.E, A.J, A.G, and M.A:** Writing – Reviewing and Editing.

## Data Availability

All the data used to conduct this manuscript are included in tables, figures, and mentioned in the manuscript, including references.

## Acknowledgments

None.

## References

1. Chioncel O, Parissis J, Mebazaa A, et al. Epidemiology, pathophysiology and contemporary management of cardiogenic shock – a position statement from the Heart Failure Association of the European Society of Cardiology. Eur J Heart Fail. 2020;22(8):1315–1341. 10.1002/ejhf.1922

2. Hochman JS, Sleeper LA, Webb JG, et al. Early Revascularization in Acute Myocardial Infarction Complicated by Cardiogenic Shock. N Engl J Med. 1999;341(9):625–634. doi:10.1056/nejm199908263410901

3. Thiele H, Zeymer U, Neumann FJ, et al. Intraaortic Balloon Support for Myocardial Infarction with Cardiogenic Shock. N Engl J Med. 2012;367(14):1287–1296. doi:10.1056/NEJMoa1208410

4. Thiele H, Zeymer U, Thelemann N, et al. Intraaortic Balloon Pump in Cardiogenic Shock Complicating Acute Myocardial Infarction: Long-Term 6-Year Outcome of the Randomized IABP-SHOCK II Trial. Circulation. 2019;139(3):395–403. doi:10.1161/circulationaha.118.038201

5. Ouweneel DM, Eriksen E, Sjauw KD, et al. Percutaneous Mechanical Circulatory Support Versus Intra-Aortic Balloon Pump in Cardiogenic Shock After Acute Myocardial Infarction. J Am Coll Cardiol. 2017;69(3):278–287. doi:10.1016/j.jacc.2016.10.022

6. Basir MB, Schreiber TL, Grines CL, et al. Effect of Early Initiation of Mechanical Circulatory Support on Survival in Cardiogenic Shock. Am J Cardiol. 2017;119(6):845–851. doi:10.1016/j.amjcard.2016.11.037

7. Tehrani BN, Truesdell AG, Sherwood MW, et al. Standardized Team-Based Care for Cardiogenic Shock. J Am Coll Cardiol. 2019;73(13):1659–1669. 10.1016/j.jacc.2018.12.084

8. Esposito ML, Kapur NK. Acute mechanical circulatory support for cardiogenic shock: the “door to support” time. F1000Res. 2017;6:737. doi:10.12688/f1000research.11150.1

9. Liang Y, Nie SP, Wang X, et al. Role of Pulmonary Embolism Response Team in patients with intermediate– and high-risk pulmonary embolism: a concise review and preliminary experience from China. J Geriatr Cardiol. 2020;17(8):510–518. doi:10.11909/j.issn.1671-5411.2020.08.005

10. Morey JR, Zhang X, Marayati NF, et al. Mobile Interventional Stroke Teams Improve Outcomes in the Early Time Window for Large Vessel Occlusion Stroke. Stroke. 2021;52(9):e527–e530. doi: doi:10.1161/STROKEAHA.121.034222

11. Taleb I, Koliopoulou AG, Tandar A, et al. Shock Team Approach in Refractory Cardiogenic Shock Requiring Short-Term Mechanical Circulatory Support. Circulation. 2019;140(1):98–100. doi: doi:10.1161/CIRCULATIONAHA.119.040654

12. Sebat F, Johnson D, Musthafa AA, et al. A Multidisciplinary Community Hospital Program for Early and Rapid Resuscitation of Shock in Nontrauma Patients. Chest. 2005;127(5):1729–1743. doi:10.1378/chest.127.5.1729

13. Hong D, Choi KH, Cho YH, et al. Multidisciplinary team approach in acute myocardial infarction patients undergoing veno-arterial extracorporeal membrane oxygenation. Ann Intensive Care. 2020;10(1):83. doi:10.1186/s13613-020-00701-8

14. Lee F, Hutson JH, Boodhwani M, et al. Multidisciplinary Code Shock Team in Cardiogenic Shock: A Canadian Centre Experience. CJC Open. 2020;2(4):249–257. doi:10.1016/j.cjco.2020.03.009

15. Papolos AI, Kenigsberg BB, Berg DD, et al. Management and Outcomes of Cardiogenic Shock in Cardiac ICUs With Versus Without Shock Teams. J Am Coll Cardiol. 2021;78(13):1309–1317. 10.1016/j.jacc.2021.07.044

16. Tehrani B, Truesdell A, Singh R, Murphy C, Saulino P. Implementation of a Cardiogenic Shock Team and Clinical Outcomes (INOVA-SHOCK Registry): Observational and Retrospective Study. JMIR Res Protoc. 2018;7(6):e160. doi:10.2196/resprot.9761

17. Cumpston M, Li T, Page MJ, et al. Updated guidance for trusted systematic reviews: a new edition of the Cochrane Handbook for Systematic Reviews of Interventions. Cochrane database Syst Rev. 2019;10:ED000142. doi:10.1002/14651858.ED000142

18. Page MJ, McKenzie JE, Bossuyt PM, et al. The PRISMA 2020 statement: an updated guideline for reporting systematic reviews. BMJ. 2021;372. doi:10.1136/bmj.n71

19. Wells GA, Wells G, Shea B, et al. The Newcastle-Ottawa Scale (NOS) for Assessing the Quality of Nonrandomised Studies in Meta-Analyses. In:; 2014.

20. Shamsrizi P, Gladstone BP, Carrara E, et al. Variation of effect estimates in the analysis of mortality and length of hospital stay in patients with infections caused by bacteria-producing extended-spectrum beta-lactamases: a systematic review and meta-analysis. BMJ Open. 2020;10(1):e030266. doi:10.1136/bmjopen-2019-030266

21. Ponikowski P, Voors AA, Anker SD, et al. 2016 ESC Guidelines for the diagnosis and treatment of acute and chronic heart failure. Eur Heart J. 2016;37(27):2129. doi:10.1093/eurheartj/ehw128

22. Menon V, White H, LeJemtel T, Webb JG, Sleeper LA, Hochman JS. The clinical profile of patients with suspected cardiogenic shock due to predominant left ventricular failure: a report from the SHOCK Trial Registry. J Am Coll Cardiol. 2000;36(3, Supplement 1):1071–1076. 10.1016/S0735-1097(00)00874-3

23. Diepen S van, Katz JN, Albert NM, et al. Contemporary Management of Cardiogenic Shock: A Scientific Statement From the American Heart Association. Circulation. 2017;136(16):e232–e268. doi:doi:10.1161/CIR.0000000000000525

24. Menon V, Slater JN, White HD, Sleeper LA, Cocke T, Hochman JS. Acute myocardial infarction complicated by systemic hypoperfusion without hypotension: report of the SHOCK trial registry. Am J Med. 2000;108(5):374–380. doi:10.1016/S0002-9343(00)00310-7

25. Goldstein JA, Barzilai B, Rosamond TL, Eisenberg PR, Jaffe AS. Determinants of hemodynamic compromise with severe right ventricular infarction. Circulation. 1990;82(2):359–368. doi:doi:10.1161/01.CIR.82.2.359

26. Moghaddam N, van Diepen S, So D, Lawler PR, Fordyce CB. Cardiogenic shock teams and centres: a contemporary review of multidisciplinary care for cardiogenic shock. ESC Hear Fail. 2021;8(2):988–998. 10.1002/ehf2.13180

27. Brusca SB, Caughron H, Njoroge JN, Cheng R, O’Brien CG, Barnett CF. The shock team: a multidisciplinary approach to early patient phenotyping and appropriate care escalation in cardiogenic shock. Curr Opin Cardiol. 2022;37(3):241–249. doi:10.1097/hco.0000000000000967

28. Doll JA, Ohman EM, Patel MR, et al. A team-based approach to patients in cardiogenic shock. Catheter Cardiovasc Interv. 2016;88(3):424–433. doi:10.1002/ccd.26297

29. Kochar A, Al-Khalidi HR, Hansen SM, et al. Delays in Primary Percutaneous Coronary Intervention in ST-Segment Elevation Myocardial Infarction Patients Presenting With Cardiogenic Shock. JACC Cardiovasc Interv. 2018;11(18):1824–1833. 10.1016/j.jcin.2018.06.030

30. Scholz KH, Maier SKG, Maier LS, et al. Impact of treatment delay on mortality in ST-segment elevation myocardial infarction (STEMI) patients presenting with and without haemodynamic instability: results from the German prospective, multicentre FITT-STEMI trial. Eur Hear J. 2018;39(13):1065–1074. doi:10.1093/eurheartj/ehy004

31. Tongers J, Sieweke JT, Kühn C, et al. Early Escalation of Mechanical Circulatory Support Stabilizes and Potentially Rescues Patients in Refractory Cardiogenic Shock. Circ Hear Fail. 2020;13(3):e005853. doi:doi:10.1161/CIRCHEARTFAILURE.118.005853

32. Pieri M, Sorrentino T, Oppizzi M, et al. The role of different mechanical circulatory support devices and their timing of implantation on myocardial damage and mid-term recovery in acute myocardial infarction related cardiogenic shock. J Interv Cardiol. 2018;31(6):717–724. doi:10.1111/joic.12569

33. Meraj PM, Doshi R, Schreiber T, Maini B, O’Neill WW. Impella 2.5 initiated prior to unprotected left main PCI in acute myocardial infarction complicated by cardiogenic shock improves early survival. J Interv Cardiol. 2017;30(3):256–263. 10.1111/joic.12377

34. Kim DH. Mechanical Circulatory Support in Cardiogenic Shock: Shock Team or Bust? Can J Cardiol. 2020;36(2):197–204. doi:10.1016/j.cjca.2019.11.001

35. Bellumkonda L, Gul B, Masri SC. Evolving Concepts in Diagnosis and Management of Cardiogenic Shock. Am J Cardiol. 2018;122(6):1104–1110. doi:10.1016/j.amjcard.2018.05.040

36. Samuels LE, Kaufman MS, Thomas MP, Holmes EC, Brockman SK, Wechsler AS. Pharmacological Criteria for Ventricular Assist Device Insertion Following Postcardiotomy Shock: Experience with the Abiomed BVS System. J Card Surg. 1999;14(4):288–293. 10.1111/j.1540-8191.1999.tb00996.x

37. Na SJ, Park TK, Lee GY, et al. Impact of a cardiac intensivist on mortality in patients with cardiogenic shock. Int J Cardiol. 2017;244:220–225. doi:10.1016/j.ijcard.2017.06.082

38. Damluji AA, Myerburg RJ, Chongthammakun V, et al. Improvements in Outcomes and Disparities of ST-Segment-Elevation Myocardial Infarction Care: The Miami-Dade County ST-Segment-Elevation Myocardial Infarction Network Project. Circ Cardiovasc Qual Outcomes. 2017;10(12). doi:10.1161/circoutcomes.117.004038

39. Vallabhajosyula S, Dunlay SM, Barsness GW, Rihal CS, Holmes Jr. DR, Prasad A. Hospital-Level Disparities in the Outcomes of Acute Myocardial Infarction With Cardiogenic Shock. Am J Cardiol. 2019;124(4):491–498. doi:10.1016/j.amjcard.2019.05.038

40. Shaefi S, O’Gara B, Kociol RD, et al. Effect of Cardiogenic Shock Hospital Volume on Mortality in Patients With Cardiogenic Shock. J Am Heart Assoc. 2015;4(1):e001462. doi:doi:10.1161/JAHA.114.001462

41. Loccoh EC, Joynt Maddox KE, Wang Y, Kazi DS, Yeh RW, Wadhera RK. Rural-Urban Disparities in Outcomes of Myocardial Infarction, Heart Failure, and Stroke in the United States. J Am Coll Cardiol. 2022;79(3):267–279. 10.1016/j.jacc.2021.10.045

42. Vallabhajosyula S, Patlolla SH, Dunlay SM, et al. Regional Variation in the Management and Outcomes of Acute Myocardial Infarction With Cardiogenic Shock in the United States. Circ Hear Fail. 2020;13(2):e006661. doi:doi:10.1161/CIRCHEARTFAILURE.119.006661

43. Baran, DA, Grines, CL, Bailey, S, et al. SCAI clinical expert consensus statement on the classification of cardiogenic shock. Catheter Cardiovasc Interv. 2019; 94: 29–37. 10.1002/ccd.28329

44. Naidu S, Baran D, Jentzer J, et al. SCAI SHOCK Stage Classification Expert Consensus Update: A Review and Incorporation of Validation Studies. J Am Coll Cardiol. 2022 Mar, 79 (9) 933–946.

45. Arrigo, M., Jessup, M., Mullens, W. et al. Acute heart failure. Nat Rev Dis Primers 6, 16 (2020). 10.1038/s41572-020-0151-7

46. Morici N, Viola G, Antolini L, Alicandro G, Dal Martello M, Sacco A, Bottiroli M, Pappalardo F, Villanova L, De Ponti L, La Vecchia C, Frigerio M, Oliva F, Fried J, Colombo P, Garan AR. Predicting survival in patients with acute decompensated heart failure complicated by cardiogenic shock. Int J Cardiol Heart Vasc. 2021 Jun 4;34:100809. doi: 10.1016/j.ijcha.2021.100809. PMID: 34141863; PMCID: PMC8188054.

